# Scoping Review of One-Dimension Statistical Parametric Mapping in Lower Limb Biomechanical Analysis

**DOI:** 10.1101/2023.08.06.23293709

**Authors:** Tomer Yona, Netanel Kamel, Galya Cohen-Eick, Inbar Ovadia, Arielle Fischer

## Abstract

**Background:** Biomechanics is crucial in enhancing sports performance and preventing injury. Traditionally, discrete point analysis is used to analyze important kinetic and kinematic data points, reducing continuous data to a single point. One-dimensional Statistical Parametric Mapping (spm1d) offers a more comprehensive approach by assessing entire movement curves instead of isolated peak values. Nevertheless, spm1d is still underutilized in various sports and sports-related injuries.

**Purpose:** To summarize the existing literature on the application of spm1d in sports biomechanics, including the kinetics and kinematics of the hip, knee, and ankle joints, as well as to identify gaps in the literature that may require further research.

**Methods:** A scoping review was conducted, searching PubMed, Embase, Web of Science, and ProQuest databases. English peer-reviewed studies using SPM to assess lower limb kinetics or kinematics in different sports or sports-related injuries were included. In contrast, reviews, meta-analyses, conference abstracts, grey literature, and studies focusing on non-kinetic or kinematic outcomes were excluded.

**Results:** The review yielded 129 papers, with an increased number of studies published in the last three years. Of these studies, 81 examined healthy individuals (63%), and 48 focused on injured populations (37%). Running (n=28), cutting (n=21), and jumping/landing (n=14) were the most common activities. The most prevalent sport-related injuries examined were anterior cruciate ligament rupture (n=21), chronic ankle instability (n=16), and hip-related pain (n=9). Research gaps include the underrepresentation of common sports and movements, small sample size, lack of studies in non-laboratory settings and varied active age groups, and absence of evaluations on the effects of protective sports gear other than shoes.

**Conclusion:** The application of spm1d in sports biomechanics demonstrates diverse uses in sports performance, injury reduction, and rehabilitation. While spm1d shows promise in improving our understanding of sports biomechanics, there are still significant gaps in the literature that present future research opportunities.

## INTRODUCTION

From enhancing performance to preventing injury, biomechanics plays a pivotal role in the field of sports and sports-related injuries.^1, 2^ Traditionally, kinetics and kinematics are collected as continuous signals and analyzed using discrete point analysis, which reduces the data to isolated points, such as the minima and maxima of the signal. However, this approach may overlook important information within the continuous signal under analysis, such as the range of a joint’s movement, ground reaction forces, and moments acting on a joint during different activities.

Statistical Parametric Mapping (SPM) has been widely used in neuroimaging as it allows us to see changes across the entire brain, not just in specific areas.^3^ This approach has advantages over simpler, point-by-point methods, which might miss these widespread changes. Pataky (2010)^4^ introduced one-dimensional statistical parametric mapping (spm1d) as a method for biomechanical analysis, enabling the assessment of entire movement curves for statistical significance. By employing spm1d, researchers can gain a more comprehensive understanding of movement biomechanics instead of solely focusing on the peak values of specific movements.

This review focuses on the biomechanics of the lower limbs due to their crucial role in most sports activities and their susceptibility to injury. Through the application of spm1d, researchers can analyze simple and complex movements of the hip, knee, and ankle joints, identifying subtle patterns that may not be evident through traditional discrete point analysis. This improved understanding can lead to enhanced performance, effective treatment approaches for sports injuries, precise strength and conditioning programs, and improved rehabilitation strategies. Furthermore, it can improve the development and design of sports equipment and protective gear.

Given the early stages of spm1d in biomechanics, this scoping review offers a unique opportunity to uncover research trajectories and address knowledge gaps. Unlike systematic reviews focusing on synthesizing conclusive evidence for specific questions, scoping reviews are better suited for exploring broad research areas.

Through this scoping review, we aim to examine the existing literature in sports medicine and sports biomechanics utilizing spm1d, describe the studies characteristics, identify knowledge gaps, and propose future research directions. This approach is particularly suitable given the wide-ranging applications of spm1d in biomechanics.^5^ Specifically, our focus will be on the lower limbs, including the kinetics and kinematics of the hip, knee, and ankle joints.

## METHODS

To comprehensively review the applications of SPM in biomechanics, we conducted a scoping review of the literature. We performed a search in PubMed, Embase, Web of Science, and ProQuest, with the following search string: “(((knee) OR (hip)) OR (ankle)) AND (statistical parametric mapping)”. The specific search strategy for each database is detailed in Appendix 1.

We included peer-reviewed studies written in English that utilized spm1d as the main outcome to assess lower limb kinetics or kinematics in different sports or common sports injuries. Studies comparing different measurement tools or assessing electromyography as the primary outcome were excluded. Additionally, studies unrelated to sports or sport-related injuries were excluded, along with reviews, meta-analyses, conference abstracts, and grey literature. There were no restrictions on the publication date. The database search was conducted in March 2023.

Two authors independently screened the titles and abstracts of the identified papers using the Rayyan online system.^6^ Subsequently, using a custom-written Excel file, two authors independently reviewed the full text of the included studies and extracted the following information from each study: Study design, number of participants, population (healthy/injured), sport played by the participants, activity and joints assessed in the study, measurement tools, and outcome measures. Any discrepancies were solved through discussion. Before commencing the review, all the authors piloted the system and the screening process and established a consensus.

This study adheres to the PRISMA Extension for Scoping Reviews (PRISMA-ScR) Checklist.^7^ The protocol for this study was registered in the Open Science Framework before initiating the review and is available online.^8^

## RESULTS

The initial search identified 1305 records. After removing duplicates, 531 papers underwent title and abstract screening. Subsequently, the full text of 132 studies was screened, resulting in one study being excluded due to a wrong primary outcome, one due to the wrong population, and one not available to the authors (Figure 1). Finally, 129 papers were included in this review, with over half published in the last three years (Figure 2). Most studies assessed healthy populations (n=81), and 48 involved injured participants. The median number of participants in the studies among healthy participants was 19 [9-90], and the median among injured participants was 27.5 [9-357].

**Figure 1.**
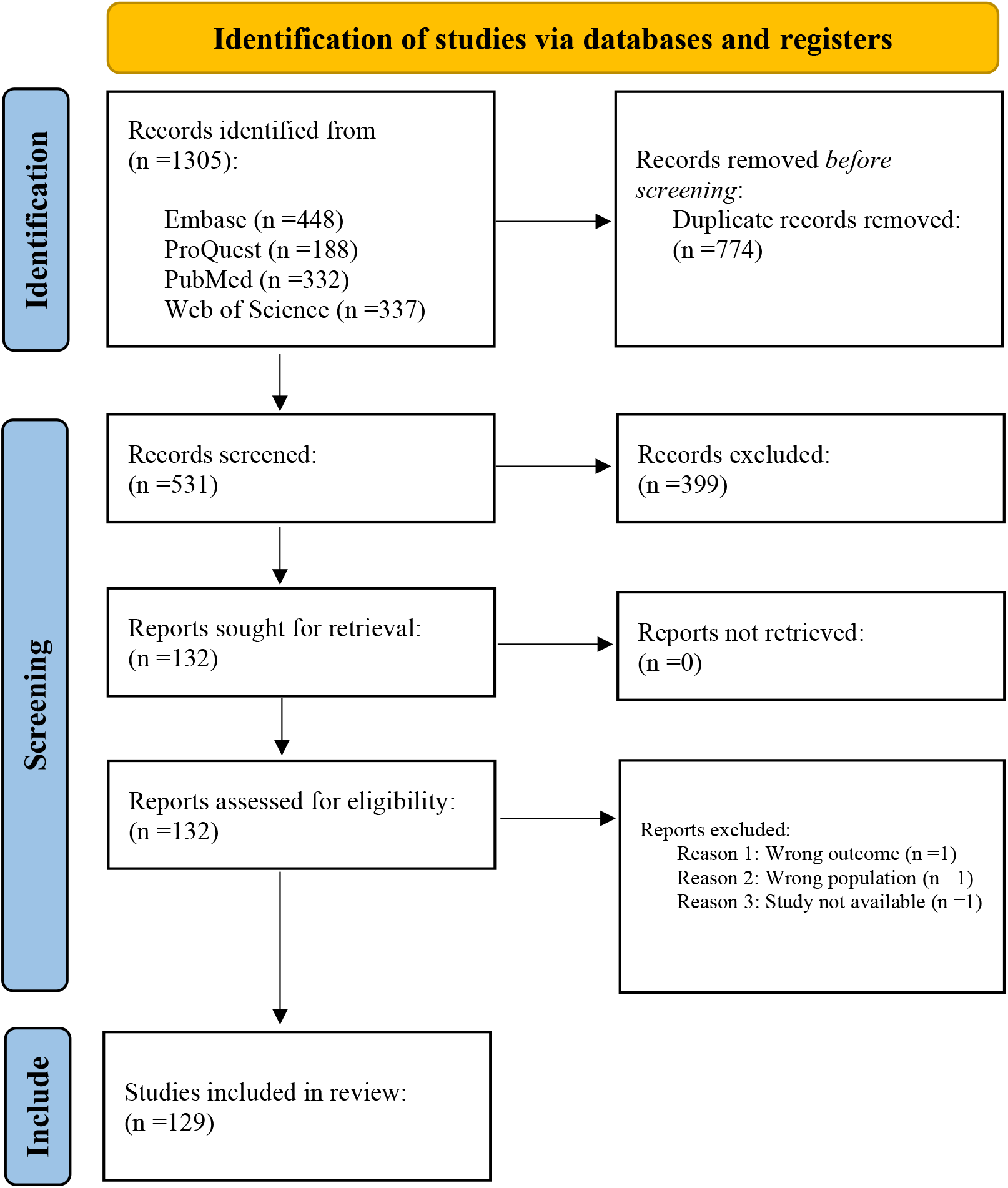
PRISMA flow diagram.

**Figure 2.**
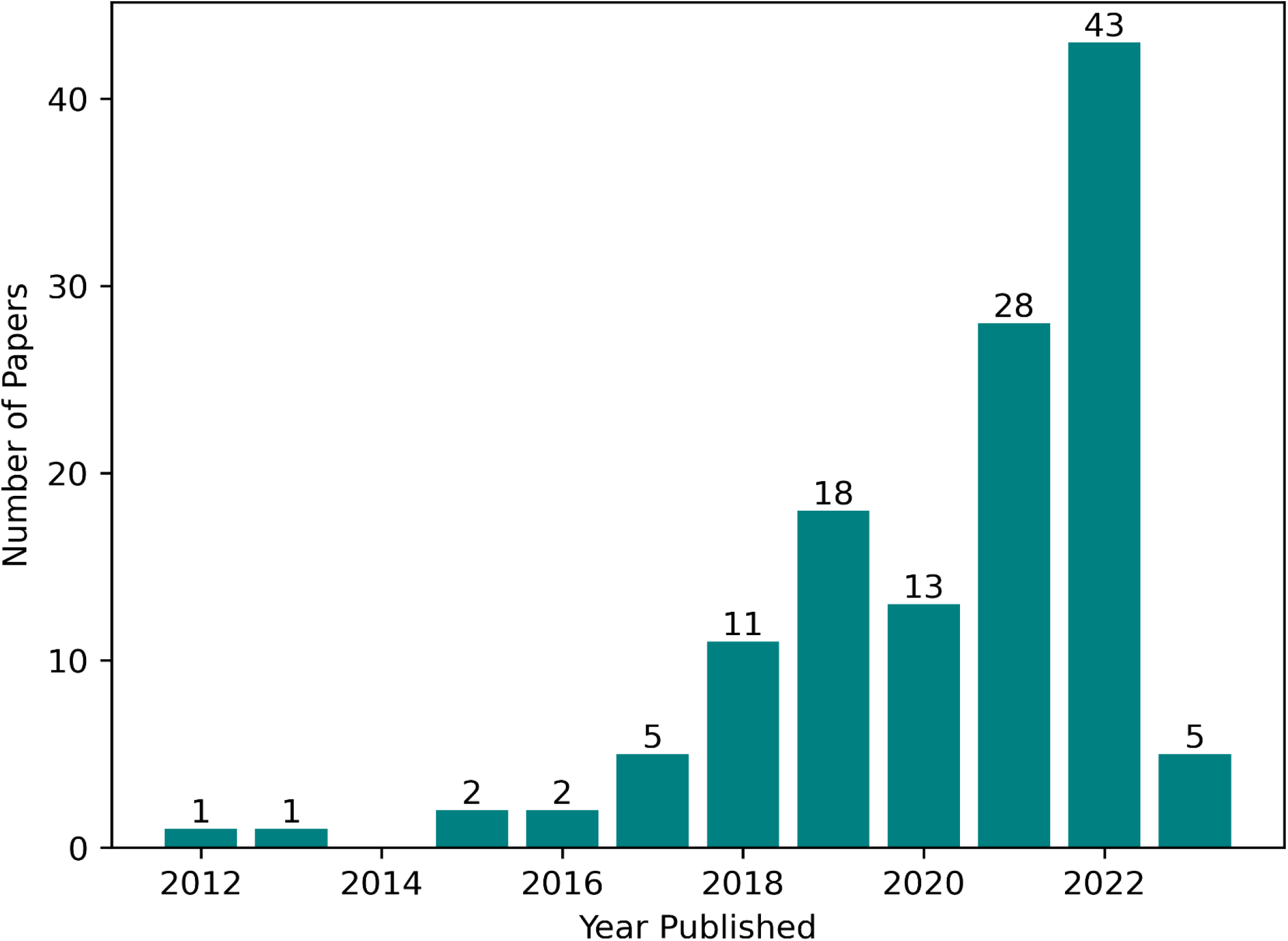
Number of Studies Utilizing spm1d for lower limb biomechanics Published per Year.

### Running

A total of 28 studies assessed running, 13 involving treadmill running and 15 surface running. Among these studies, ten focused on the effects of different shoes on running biomechanics,^9–18^ three studies examined specific physical interventions,^19–21^ three studies assessed various gait modifications,^22–24^ three evaluated foot biomechanics,^25–27^ two explored the effects of fatigue,^28, 29^ and two investigated studies the effects of sex.^30, 31^ Additionally, single studies examined the effects of different surfaces,^32^ hamstring flexibility,^33^pertubations,^34^ running with different loads,^35^ and the differences between transition running and isolated running in triathlon Table 1).^36^ The median number of participants was 17.5 [9-87].

**Table 1.** SPM1d in Assessing Running Activities.

| Participants Details |  |  |  | SPM Assessment |  |  |  |
| --- | --- | --- | --- | --- | --- | --- | --- |
| Study | Number Of Participants | Sport participation | Activity | Joints | Measurement Tools | Kinetic/ Kinematic | SPM/ Discreet |
| Shamsoddini (2022)<sup>14</sup> | 17 | Physically active | Running | Hip, Knee, Ankle | Qualisys + Kistler | Kinematics + Kinetics | SPM |
| Zhang (2022)<sup>18</sup> | 13 | Running | Running | Pelvis, Hip, Knee | Motion Analysis + Kistler | Kinematics + Kinetics | SPM+ Discreet |
| Chen (2022)<sup>10</sup> | 10 | Running | Running | Hip, Knee, Ankle | Vicon + AMTI | Kinematics + Kinetics | SPM+ Discreet |
| Fu (2022)<sup>12</sup> | 15 | Running | Running | Hip, Knee, Ankle | Vicon + AMTI | Kinematics + Kinetics | SPM+ Discreet |
| Besson (2019)<sup>9</sup> | 15 | Running | Running | Hip, Knee, Ankle | Motion Analysis + Kistler | Kinematics + Kinetics | SPM+ Discreet |
| Sinclair (2018)<sup>15</sup> | 15 | Running | Running | Knee | Qualisys + Kistler | Kinetics | SPM |
| Tam (2017)<sup>17</sup> | 50 | Running | Running | Knee, Ankle | Vicon + AMTI | Kinematics + Kinetics | SPM |
| Sinclair (2021)<sup>16</sup> | 13 | Running | Running | Hip, Knee, Ankle | Qualisys + Kistler | Kinematics + Kinetics | SPM |
| Costa (2021)<sup>11</sup> | 16 | Running | Running- Treadmill | Hip, Knee, Ankle | Qualisys + Bertec | Kinematics + Kinetics | SPM+ Discreet |
| Nüesch (2019)<sup>13</sup> | 19 | Running | Running- Treadmill | Hip, Knee, Ankle | RehaGait + Zebris | Kinematics + Kinetics | SPM |
| Nishida (2022)<sup>30</sup> | 19 | Collegiate athletes | Running- Treadmill | Knee | XRAY + Bertec | Kinematics | SPM+ Discreet |
| Takabayashi (2019)<sup>31</sup> | 22 | Running, Jogging | Running- Treadmill | Ankle | Vicon | Kinematics | SPM |
| Gao (2022)<sup>28</sup> | 18 | Running | Running- Treadmill | Hip, Knee, Ankle | Vicon + Kistler | Kinematics | SPM |
| Möhler (2022)<sup>29</sup> | 14 | Running | Running- Treadmill | Hip, Knee, Ankle, CM | Vicon | Kinematics | SPM+ Discreet |
| Matias (2022)<sup>20</sup> | 87 | Running | Running- Treadmill | Ankle, foot | Vicon + AMTI | Kinematics | SPM |
| Trowell (2022)<sup>21</sup> | 28 | Running | Running- Treadmill | Hip, Knee, Ankle | Vicon + Kistler | Kinematics + Kinetics | SPM+ Discreet |
| Masters (2018)<sup>19</sup> | 23 | Running | Running | Hip, Knee | Vicon + AMTI | Kinematics | SPM+ Discreet |
| Bennett (2021)<sup>23</sup> | 19 | Running | Running | Hip, Knee, Ankle | Qualisys + AMTI | Kinetics | SPM |
| Alizadeh (2019)<sup>22</sup> | 34 | Soccer | Running - Treadmill | Hip, Knee | Vicon, Diers Formetric 4D | Kinematics | SPM+ Discreet |
| Mei (2019)<sup>24</sup> | 20 | Running | Running- Treadmill | Hip, Knee, Ankle | Vicon, Diers Formetric 4D | Kinematics + Kinetics | SPM+ Discreet |
| Deschamps (2022)<sup>25</sup> | 12 | Recreationally active | Running | Ankle | Vicon + AMTI | Kinematics + Kinetics | SPM |
| Glassbrook (2020)<sup>26</sup> | 16 | Recreationally active | Running- Treadmill | Ankle | iMeasureU | Kinematics | SPM+ Discreet |
| Judson (2019)<sup>27</sup> | 9 | Running, Sprint | Sprint | Foot | Motion Analysis + Kistler | Kinetics | SPM |
| Zhou (2023)<sup>32</sup> | 30 | Running | Running | Hip, Knee, Ankle | Vicon + Kistler | Kinematics + Kinetics | SPM |
| Garcia (2022)<sup>33</sup> | 34 | Running | Running- Treadmill | Pelvis, Hip, Knee | Motion Analysis | Kinematics | SPM+ Discreet |
| Khajooei (2022)<sup>34</sup> | 13 | Running | Running- Treadmill | Hip, Knee, Ankle | Vicon | Kinematics | SPM |
| Fraeulin (2021)<sup>36</sup> | 16 | Running, Triathlon | Running | Hip, Knee, Ankle | XSENS | Kinematics | SPM+ Discreet |
| Liew (2016)<sup>35</sup> | 31 | Running | Running | Hip, Knee, Ankle | Vicon + AMTI | Kinematics + Kinetics | SPM |

### Cutting

A total of 21 studies assessed cutting/change of direction activities. Among them, five studies evaluated the effect of foot-strike and movement patterns on cutting maneuvers,^37–41^ four focused on the influence of anticipation and uncertainty,^42–45^ four assessed the influence of external factors such as fatigue,^46, 47^ footwear^48^ and surface,^49^ and two looked at the influence of training and movement strategies.^50, 51^ Single studies evaluated the effects of sex,^52^ speed,^53^ lab versus field,^54^ limb differences,^55^ cutting on a softball base compared to a flat surface,^56^ and the joint contact forces of the medial versus the lateral tibiofemoral joint (table 2).^57^ The median number of participants was 24 [12-50].

**Table 2.** SPM1d in Assessing Cutting Activities.

| Participants Details |  |  |  | SPM Assessment |  |  |  |
| --- | --- | --- | --- | --- | --- | --- | --- |
| Study | Number Of Participants | Sport participation | Activity | Joints | Measurement Tools | Kinetic/ Kinematic | SPM/ Discreet |
| Uno (2022)<sup>41</sup> | 23 | Team sport | Cutting | Knee, GRF, CM | OptiTrack + AMTI | Kinematics + Kinetics | SPM |
| Ogasawara (2021)<sup>39</sup> | 25 | Handball | Cutting | Hip, Knee | OptiTrack + Kistler | Kinematics + Kinetics | SPM+ Discreet |
| Ogasawara (2020)<sup>38</sup> | 24 | Handball | Cutting | Knee, Ankle | OptiTrack + Kistler | Kinetics | SPM |
| Peel (2022)<sup>40</sup> | 24 | Recreationally active | Cutting | Knee, Ankle | Vicon + AMTI | Kinematics + Kinetics | SPM+ Discreet |
| David (2017)<sup>37</sup> | 50 | N/A | Cutting | Knee | Vicon + Kistler | Kinematics + Kinetics | SPM+ Discreet |
| Whyte (2018)<sup>45</sup> | 28 | Gaelic football | Cutting | Hip, Knee, Ankle | Vicon + AMTI | Kinematics + Kinetics | SPM |
| Whyte (2018)<sup>44</sup> | 28 | Gaelic football | Side Cutting | Hip, Knee, Ankle | Vicon + AMTI | Kinematics + Kinetics | SPM |
| Bedo (2021)<sup>42</sup> | 31 | Handball | Change of direction | Knee | Vicon + AMTI | Kinetics | SPM+ Discreet |
| Dutailis (2021)<sup>43</sup> | 19 | Recreationally active | Sidestep cutting | Hip, Knee, Ankle | Vicon + AMTI | Kinematics | SPM |
| Schroeder (2021)<sup>56</sup> | 16 | Softball | Change of direction | Hip, Knee, Ankle | Vicon + AMTI | Kinematics + Kinetics | SPM+ Discreet |
| Cassiolas (2022)<sup>57</sup> | 31 | Recreational and elite athletes | Change of direction | Knee | Vicon + AMTI | Kinetics | SPM+ Discreet |
| Thomas (2022)<sup>55</sup> | 14 | Soccer | Change of direction | Hip, Knee, Ankle | Qualisys + AMTI | Kinematics + Kinetics | SPM |
| Di Paolo (2023)<sup>54</sup> | 28 | Soccer | Cutting | Hip, Knee, Ankle | XSENS | Kinematics | SPM |
| Whyte (2018)<sup>51</sup> | 31 | Soccer | Cutting | Hip, Knee, Ankle | Vicon + AMTI | Kinematics + Kinetics | SPM |
| Sankey (2020)<sup>50</sup> | 20 | Soccer | Side cutting | Hip, Knee, Ankle | Vicon + Kistler | Kinematics + Kinetics | SPM |
| Zago (2021)<sup>47</sup> | 20 | Soccer | Change of direction | Hip, Knee, Ankle | BTS Bioengineering + AMTI | Kinematics + Kinetics | SPM+ Discreet |
| Sinclair (2020)<sup>49</sup> | 20 | Recreational athletes | Change of direction | Knee | Qualisys + Kistler | Kinematics + Kinetics | SPM+ Discreet |
| Bedo (2022)<sup>46</sup> | 20 | Handball | Side Cutting | Hip, Knee | Vicon + Bertec | Kinematics | SPM |
| Liu (2022)<sup>48</sup> | 12 | Basketball | Cutting | Ankle | Vicon + AMTI | Kinematics + Kinetics | SPM |
| Vanrenterghem (2012)<sup>53</sup> | 14 | Dynamic sporting activity | Cutting | Knee | Qualisys + Kistler | Kinematics + Kinetics | SPM+ Discreet |
| Weinhandl (2021)<sup>52</sup> | 38 | Recreationally active | Cutting | Hip, GRF | Vicon + Bertec | Kinematics + Kinetics | SPM |

### Jumping and Landing

Fourteen studies investigated jumping or landing tasks. Among these, four examined single-leg vs. double-leg landings and landings direction,^58–61^ and two evaluated the effects of different interventions: the effects of a video task on volleyball jump,^62^ and the effects of a new shoe on drop-landing.^63^ Two studies explored the effects of limb dominancy,^64, 65^ and another two explored the difference between males and females.^30, 52^ Single studies investigated the effects of fatigue,^46^ anticipation,^66^ different foot areas during landing,^67^ and different landing biomechanics among different jumping athletes (Table 3).^68^ The median number of participants was 20 [9-90].

**Table 3.**
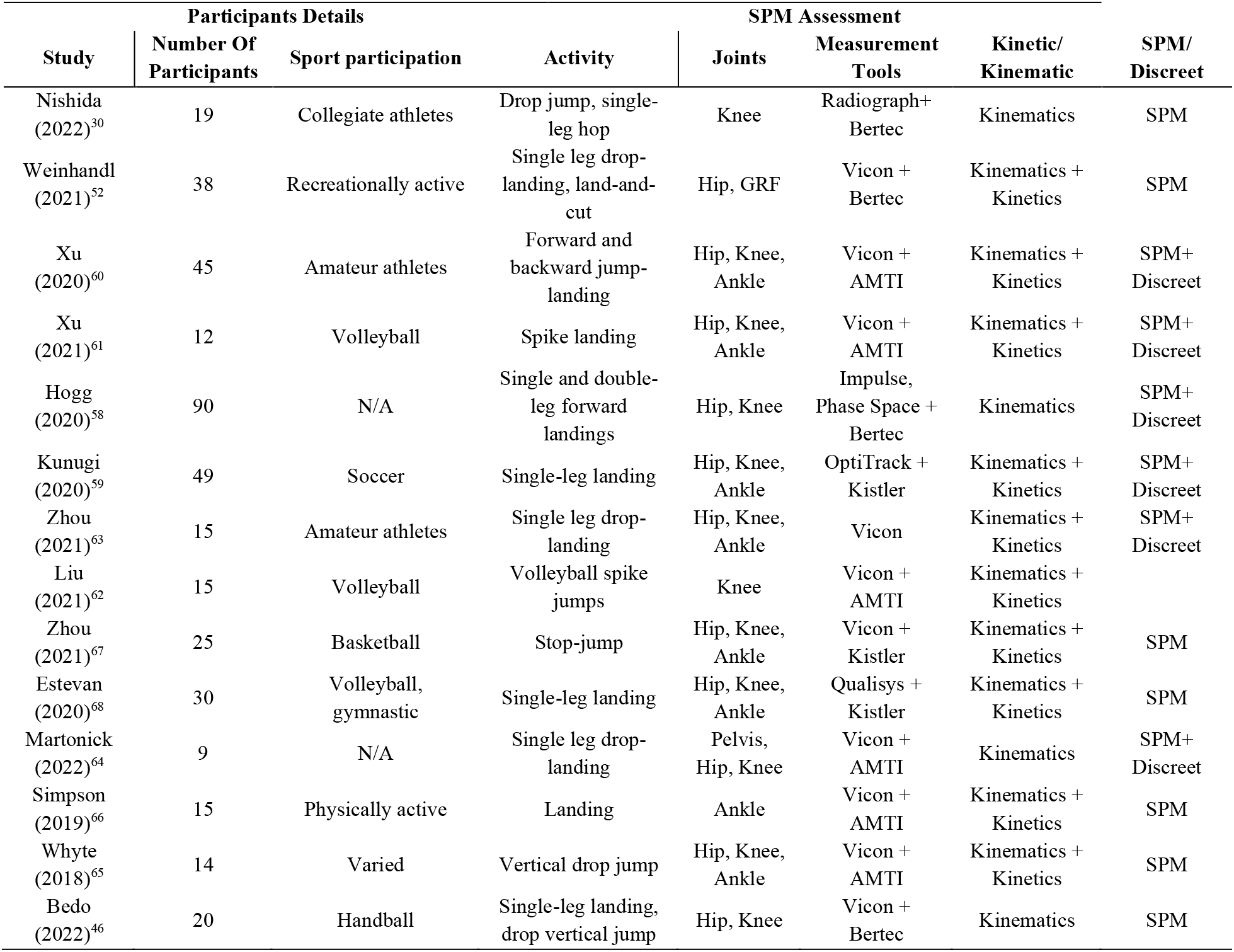
SPM1d in Assessing Jumping and Landing Activities.

### Squatting

Six studies focused on squatting, with five assessing the traditional back squat.^69–73^ One study examined a lunge squat,^74^ and one evaluated a half squat.^71^ Three studies investigated different load conditions,^69, 72, 74^ squat depth,^71^ and heel height^73^ on the biomechanics of the lower limbs. Additionally, one study evaluated the inter-individual and intra-individual variability during a squat (Table 4).^70^ The median number of participants was 15 [9-20].

**Table 4.** SPM1d in Squatting, Isokinetic and Other Activities.

| Study | Participants Details |  | Activity | Joints | SPM Assessment |  |  |
| --- | --- | --- | --- | --- | --- | --- | --- |
|  | Number Of Participants | Sport participation |  |  | Measurement Tools | Kinetic/ Kinematic | SPM/ Discreet |
| Kipp (2022)<sup>69</sup> | 9 | Track and field athletes | Squat | Hip, Knee, Ankle | Vicon + AMTI | Kinematics + Kinetics | SPM |
| Maddox (2021)<sup>72</sup> | 20 | Resistance-trained | Squat | Hip, Knee, Ankle | Vicon + Bertec | Kinematics + Kinetics | SPM+ Discreet |
| Gao (2022)<sup>74</sup> | 14 | Squat | Lunge Squat | Hip, Knee, Ankle | Vicon + Kistler | Kinematics + Kinetics | SPM, 2-Way Anova |
| Li (2021)<sup>71</sup> | 16 | Squat | Squat, half-squat | Hip, Knee, Ankle | Vicon + AMTI | Kinematics + Kinetics | SPM+ Discreet |
| Sayers (2020)<sup>73</sup> | 20 | Resistance-trained | Squat | Hip, Knee, Ankle | Vicon + Kistler | Kinematics + Kinetics | SPM+ Discreet |
| Kristiansen (2019)<sup>70</sup> | 10 | Resistance-trained | Squat | Hip, Knee, Ankle | Qualisys | Kinematics | SPM+ Discreet |
| Alhammoud (2019)<sup>76</sup> | 28 | Other, Ski | Isokinetic device | Knee | CSMi | Kinetics | SPM, Discreet |
| Oranchuk (2021)<sup>77</sup> | 14 | Resistance-trained | Isokinetic device | Knee | CSMi | Kinetics | SPM, Discreet |
| Zhang (2021)<sup>75</sup> | 17 | Soccer | Isokinetic device | Knee | CON-TREX® | Kinetics | SPM, Discreet |
| Galindo-Martínez (2021)<sup>78</sup> | 23 | Cycling | Cycling | Hip, Knee, Ankle | Vicon | Kinematics | SPM |
| Bini (2021)<sup>79</sup> | 10 | Cycling | Cycling | Hip, Knee | XSENS | Kinematics + Kinetics | SPM |
| Park (2022)<sup>80</sup> | 10 | Cycling, Stand-up cycling | Stand-up cycling | Hip, Knee, Ankle | Motion Analysis + load cell | Kinematics + Kinetics | SPM+ Discreet |
| Bertozzi (2022)<sup>84</sup> | 11 | Kayak | Kayak | Hip, Knee, Ankle | BTS bioengineering | Kinematics | SPM |
| Klitgaard (2021)<sup>85</sup> | 11 | Kayak- ergometer | Kayak | Knee | XSENS | Kinematics | SPM |
| Callaghan (2019)<sup>87</sup> | 9 | Cricket | Cricket | Knee | XSENS + Kistler | Kinematics + Kinetics | SPM+ Discreet |
| Wallace (2021)<sup>88</sup> | 13 | Dance | Dance | Hip, Knee, Ankle | Vicon + AMTI | Kinematics + Kinetics | SPM+ Discreet |
| Bissas (2022)<sup>86</sup> | 16 | Hurdling | Hurdling | Hip, Knee, Ankle, CM | Sony + SIMI | Kinematics | SPM+ Discreet |
| Augustus (2017)<sup>82</sup> | 9 | Soccer | Instep kick | Hip, Knee | Vicon + Kistler | Kinematics + Kinetics | SPM |
| Iitake (2022)<sup>81</sup> | 14 | Soccer | Instep kicking | Hip, Knee | Vicon | Kinematics + Kinetics | SPM |
| Atack (2019)<sup>83</sup> | 33 | Rugby | Place kicking | Hip, Knee | Vicon + Kistler | Kinematics + Kinetics | SPM+ Discreet |
| Schroeder (2020)<sup>89</sup> | 16 | Recreationally active | Sidestep | Hip, Knee, Ankle | Vicon + AMTI | Kinematics + Kinetics | SPM+ Discreet |

### Isokinetic

Three studies assessed the quadriceps or hamstrings muscle force curve produced by an isokinetic machine; They examined the influence of fatigue^75^ and sex^76^ on force production, as well as the kinetics and kinematics of eccentric, quasi-isometric loading (Table 4).^77^ The median number of participants was 17 [14-28].

### Other Activities

A total of 12 studies assessed other sports activities. Three studies focused on different aspects of cycling: the first examined the impact of fatigue,^78^ the second investigated the influence of saddle height,^79^ and the third evaluated the biomechanical implications of crank length.^80^ Three studies evaluated kicking; The first study compared the differences between male and female soccer players.^81^ The second study examined the effects of technique refinement intervention on soccer kick performance.^82^ The third study describes the biomechanical differences between rugby kickers with performance outcomes.^83^

Two studies were conducted on kayaking: The first evaluated the effects of fatigue on kayaking,^84^ while the second compared on-ergometer and on-water kayaking movements.^85^

Lastly, one study evaluated sex differences in hurdling,^86^ while another investigated how the duration of a cricket match impacts lower limb biomechanics.^87^ Additional study explored the correlations between the hip-ankle and knee-ankle movements during Irish dancing.^88^ Another study assessed how different types of unanticipated stimuli affect the biomechanics of sidestepping (Table 4).^89^ The median number of participants was 12 [9-33].

### Anterior Cruciate Ligament Injuries

A total of 21 studies analyzed the movements of individuals after an ACL injury. Two studies assessed walking on a treadmill,^90, 91^ one assessed walking overground^92^, and two evaluated the effects of functional resistance training on gait asymmetries.^93, 94^ Two studies assessed stairs ambulation: one assessed stairs descent and the other ascent. Both studies used a custom-built 3-step staircase.^92, 95^

Six studies looked at different jumping/landing activities, including two that assessed the effects of a knee sleeve on jumping and landing biomechanics,^96, 97^ one that assessed the effect of fatigue,^98^ and three that reported on between-limb differences.^99–101^ Lastly, three studies evaluated cutting activities.^101–103^ Additional seven studies have used isokinetic devices to evaluate the isokinetic strength of the quadriceps and hamstrings (Table 5).^104–110^ The median number of participants was 30 [12-357].

**Table 5.**
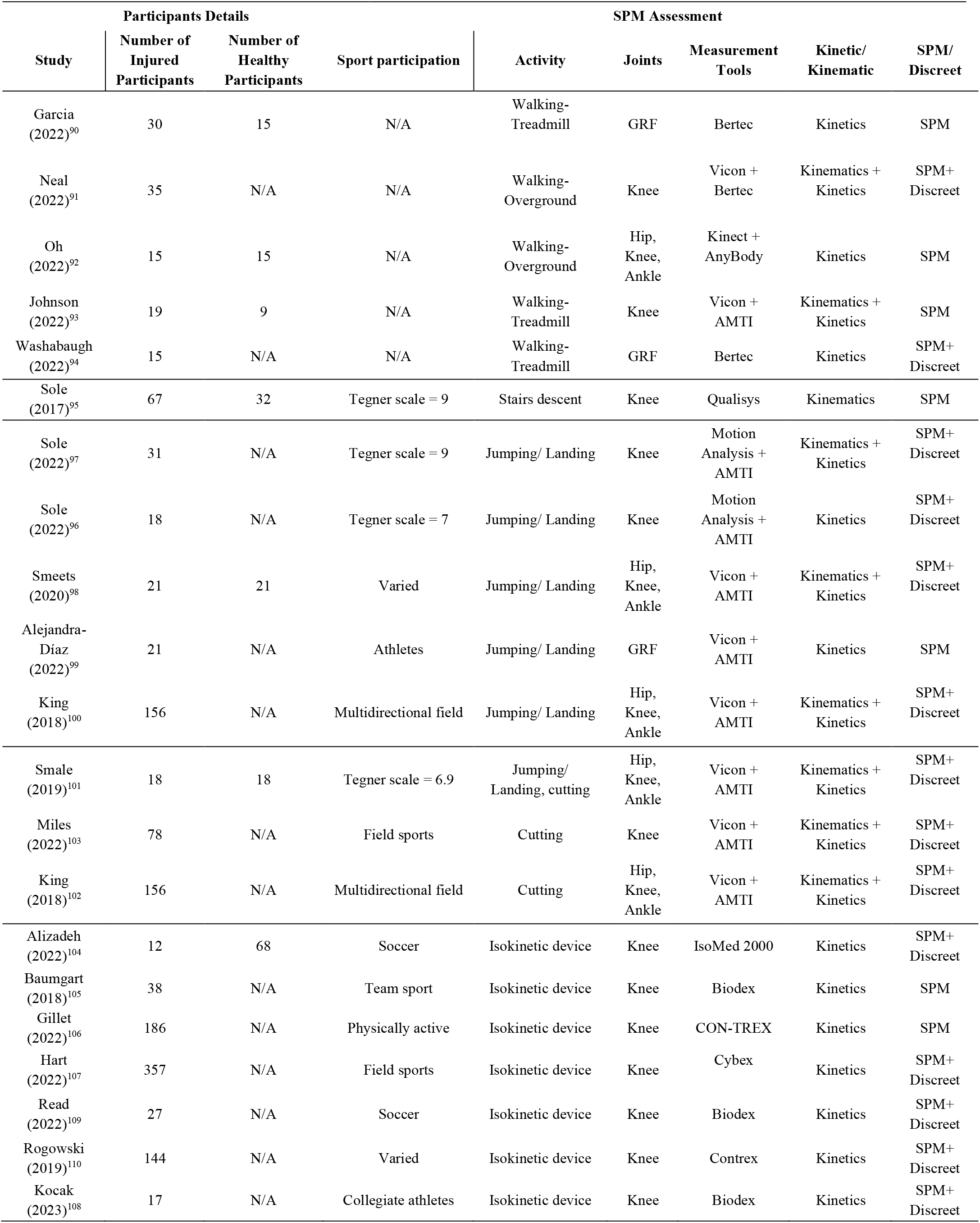
SPM1d Uses in population after an ACL Injuries.

| Study | Participants Details |  |  | SPM Assessment |  |  |  |  |
| --- | --- | --- | --- | --- | --- | --- | --- | --- |
|  | Number of Injured Participants | Number of Healthy Participants | Sport participation | Activity | Joints | Measurement Tools | Kinetic/ Kinematic | SPM/ Discreet |
| Garcia (2022)<sup>90</sup> | 30 | 15 | N/A | Walking-Treadmill | GRF | Bertec | Kinetics | SPM |
| Neal (2022)<sup>91</sup> | 35 | N/A | N/A | Walking-Overground | Knee | Vicon + Bertec | Kinematics + Kinetics | SPM+ Discreet |
| Oh (2022)<sup>92</sup> | 15 | 15 | N/A | Walking-Overground | Hip, Knee, Ankle | Kinect + AnyBody | Kinetics | SPM |
| Johnson (2022)<sup>93</sup> | 19 | 9 | N/A | Walking-Treadmill | Knee | Vicon + AMTI | Kinematics + Kinetics | SPM |
| Washabaugh (2022)<sup>94</sup> | 15 | N/A | N/A | Walking-Treadmill | GRF | Bertec | Kinetics | SPM+ Discreet |
| Sole (2017)<sup>95</sup> | 67 | 32 | Tegner scale = 9 | Stairs descent | Knee | Qualisys | Kinematics | SPM |
| Sole (2022)<sup>97</sup> | 31 | N/A | Tegner scale = 9 | Jumping/ Landing | Knee | Motion Analysis + AMTI | Kinematics + Kinetics | SPM+ Discreet |
| Sole (2022)<sup>96</sup> | 18 | N/A | Tegner scale = 7 | Jumping/ Landing | Knee | Motion Analysis + AMTI | Kinetics | SPM+ Discreet |
| Smeets (2020)<sup>98</sup> | 21 | 21 | Varied | Jumping/ Landing | Hip, Knee, Ankle | Vicon + AMTI | Kinematics + Kinetics | SPM+ Discreet |
| Alejandra-Díaz (2022)<sup>99</sup> | 21 | N/A | Athletes | Jumping/ Landing | GRF | Vicon + AMTI | Kinetics | SPM |
| King (2018)<sup>100</sup> | 156 | N/A | Multidirectional field | Jumping/ Landing | Hip, Knee, Ankle | Vicon + AMTI | Kinematics + Kinetics | SPM+ Discreet |
| Smale (2019)<sup>101</sup> | 18 | 18 | Tegner scale = 6.9 | Jumping/ Landing, cutting | Hip, Knee, Ankle | Vicon + AMTI | Kinematics + Kinetics | SPM+ Discreet |
| Miles (2022)<sup>103</sup> | 78 | N/A | Field sports | Cutting | Knee | Vicon + AMTI | Kinematics + Kinetics | SPM+ Discreet |
| King (2018)<sup>102</sup> | 156 | N/A | Multidirectional field | Cutting | Hip, Knee, Ankle | Vicon + AMTI | Kinematics + Kinetics | SPM+ Discreet |
| Alizadeh (2022)<sup>104</sup> | 12 | 68 | Soccer | Isokinetic device | Knee | IsoMed 2000 | Kinetics | SPM+ Discreet |
| Baumgart (2018)<sup>105</sup> | 38 | N/A | Team sport | Isokinetic device | Knee | Biodex | Kinetics | SPM |
| Gillet (2022)<sup>106</sup> | 186 | N/A | Physically active | Isokinetic device | Knee | CON-TREX | Kinetics | SPM |
| Hart (2022)<sup>107</sup> | 357 | N/A | Field sports | Isokinetic device | Knee | Cybex | Kinetics | SPM+ Discreet |
| Read (2022)<sup>109</sup> | 27 | N/A | Soccer | Isokinetic device | Knee | Biodex | Kinetics | SPM+ Discreet |
| Rogowski (2019)<sup>110</sup> | 144 | N/A | Varied | Isokinetic device | Knee | Contrex | Kinetics | SPM+ Discreet |
| Kocak (2023)<sup>108</sup> | 17 | N/A | Collegiate athletes | Isokinetic device | Knee | Biodex | Kinetics | SPM+ Discreet |

### Ankle Injuries

Sixteen studies evaluated people with chronic ankle instability (CAI). Three studies assessed overground walking,^111–113^ two assessed treadmill walking,^114, 115^ and one study examined the effects of taping on the foot and ankle biomechanics.^116^ Additionally, five studies focused on running biomechanics, with three being observational studies,^113, 117, 118^ and two evaluating the effects of taping.^119, 120^ Furthermore, five studies evaluated jumping/landing activities, with two assessing the effects of taping on the ankle and foot,^121, 122^ and three being observational.^123–125^ Lastly, a single study examined the kinematics of cutting (Table 6).^126^ The median number of participants was 19.5 [13-66].

**Table 6.** SPM1d in populations with ankle injuries.

| Participants Details |  |  |  | SPM Assessment |  |  |  |  |
| --- | --- | --- | --- | --- | --- | --- | --- | --- |
| Study | Number of Injured Participants | Number of Healthy Participants | Sport participation | Activity | Joints | Measurement Tools | Kinetic/ Kinematic | SPM/ Discreet |
| Ridder (2013)<sup>113</sup> | 53 | 24 | Recreationally active | Walking-Overground | Ankle | Qualisys + AMTI | Kinematics | SPM+ Discreet |
| Northeast (2018)<sup>112</sup> | 18 | 18 | Team sports | Walking-Overground | Hip, Knee, Ankle | Motion Analysis | Kinematics | SPM+ Discreet |
| Moisan (2020)<sup>111</sup> | 21 | 21 | N/A | Walking-Overground | Knee, Ankle | Optotrak + Bertec | Kinematics + Kinetics | SPM |
| Fraser (2019)<sup>114</sup> | 58 | 22 | Recreationally active | Walking-Treadmill | Ankle | Ascension Technologies + Bertec | Kinematics | SPM+ Discreet |
| Koldenhoven (2019)<sup>115</sup> | 18 | 18 | Physically active | Walking-Treadmill | Hip, Knee, Ankle | Vicon + Bertec | Kinematics + Kinetics | SPM+ Discreet |
| Dingenen (2017)<sup>116</sup> | 15 | 12 | Recreationally active | Walking-Overground | Ankle | Vicon + AMTI | Kinematics | SPM |
| Koldenhoven (2022)<sup>117</sup> | 13 | 13 | Recreationally active | Running-Treadmill | Hip, Knee, Ankle | Vicon + Bertec | Kinematics + Kinetics | SPM+ Discreet |
| Wanner (2019)<sup>118</sup> | 32 | N/A | Athletes | Running-Treadmill | Ankle | Qualisys + Bertec | Kinematics | SPM |
| Deschamps (2016)<sup>119</sup> | 15 | 12 | Recreationally active | Running-Overground | Ankle | Vicon + AMTI | Kinematics | SPM |
| Deschamps (2018)<sup>120</sup> | 15 | 12 | N/A | Running-Overground | Ankle | Vicon + AMTI | Kinematics | SPM+ Discreet |
| Agres (2019)<sup>121</sup> | 16 | N/A | Participation in sports | Jumping/Landing | Knee, Ankle | Vicon | Kinematics | SPM |
| Ridder (2020)<sup>122</sup> | 28 | N/A | N/A | Jumping/Landing | Ankle | Qualisys + AMTI | Kinematics | SPM+ Discreet |
| Ridder (2015)<sup>124</sup> | 28 | 28 | Recreationally active | Jumping/Landing | Hip, Knee, Ankle | Qualisys + AMTI | Kinematics | SPM |
| Ridder (2015)<sup>125</sup> | 58 | 36 | N/A | Jumping/Landing | Ankle | Qualisys + AMTI | Kinematics | SPM |
| Kawahara (2022)<sup>123</sup> | 18 | 18 | Varied | Jumping/Landing | Hip, Knee, Ankle | Motion Analysis + Kistler | Kinematics | SPM+ Discreet |
| Kunugi (2020)<sup>126</sup> | 66 | N/A | Soccer | Cutting | Hip, Knee, Ankle | OptiTrack + Kistler | Kinematics | SPM |

### Hip Related Pain

Nine studies examined people with hip related pain. Among these studies, two focused on overground walking,^127, 128^ and one assessed walking on a treadmill.^129^ Another two studies investigated the effects of exercise programs on walking biomechanics.^130, 131^ Furthermore, one study examined overground running,^132^ while another explored stair ambulation before and after hip osteochondroplasty and labral-chondral debridement.^133^ In addition, two studies assessed jumping/landing activities,^127, 134^ and two additional studies evaluated the biomechanics of squatting. One of these studies was observational,^135^ while the other assessed the effects of a targeted exercise program (Table 7).^131^ The median number of participants was 36 [9-88].

**Table 7.**
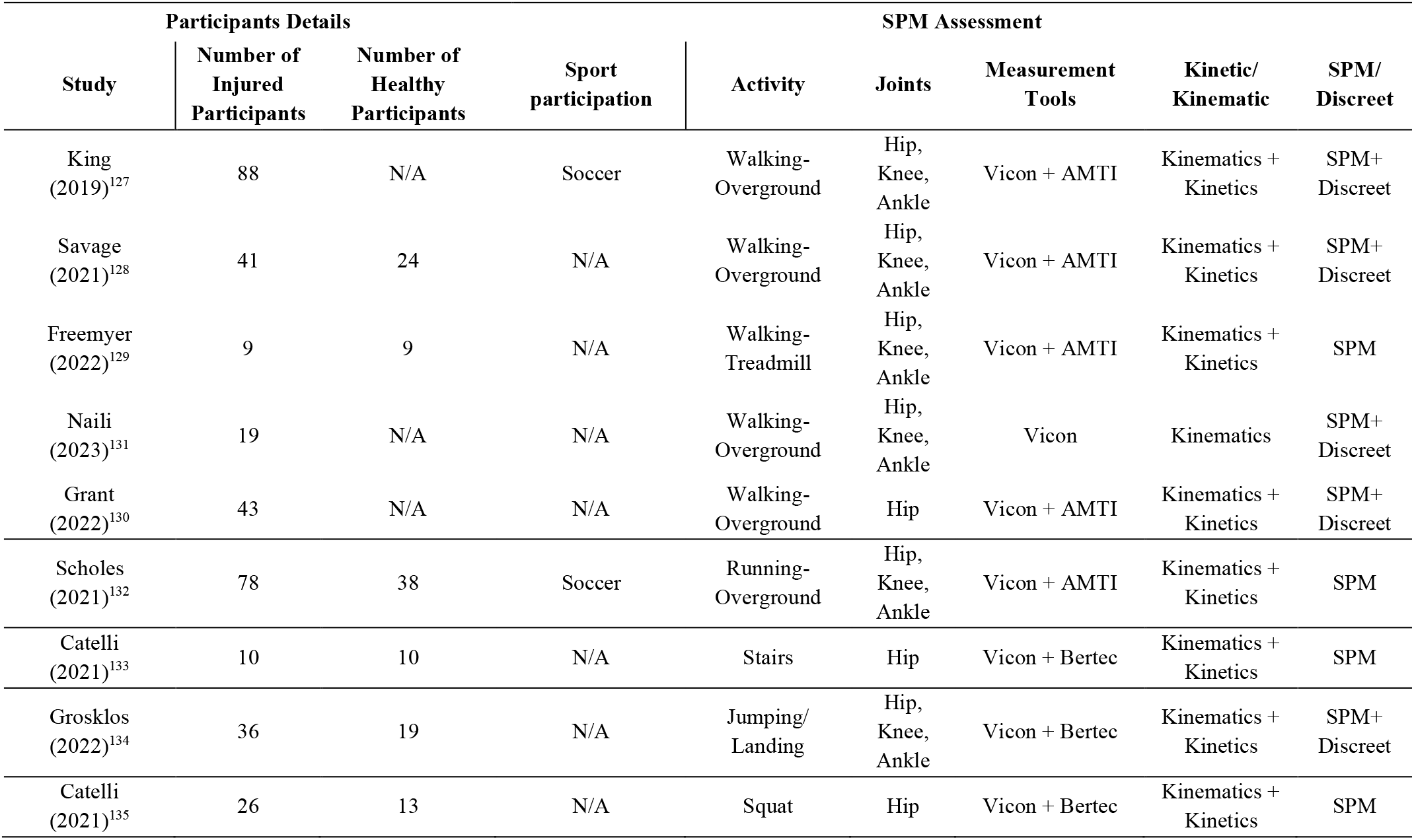
SPM1d in populations with hip related pain.

### Other Sport-Related Injuries

Three studies assessed other sport-related injuries. One investigated the effects of patellofemoral pain during squatting and walking overground.^136^ Additional two studies evaluated the impact of a hamstring injury on strength using an isokinetic device (Appendix 2).^104, 137^ The median number of participants for the above studies was 31 [25-56].

## DISCUSSION

Our systematic scoping review aimed to provide an overview of the sports and sport-injuries literature regarding using spm1d in lower limb biomechanics and identify possible research gaps. Our review identified 129 studies, 76 (59%) published in the last three years. Among the included studies, 81 (63%) focused on healthy individuals, while 48 (37%) examined injured individuals. Additionally, we found that while specific movements and sports are common in the spm1d literature, others are considerably lacking. Additionally, the median sample size of the studies was low. Next, most studies were conducted only in a laboratory setting and on young adults, ignoring other active age groups. Lastly, while some studies assessed the effects of different shoes, we found no studies assessing any protective gear.

The increasing use of spm1d over the last three years suggests a growing recognition of this method’s potential in sports biomechanics. However, the relatively low number of papers utilizing spm1d also indicated existing knowledge gaps. Furthermore, there is a lack of standardization of reporting spm1d only or together with discreet analysis; This could be attributed to researchers’ limited familiarity with spm1d and the requirement of basic coding skills, as spm1d is primarily used with MATLAB or Python.

Furthermore, the median sample size of the studies included in the analysis was relatively low, with 19 for studies involving healthy participants and 28 for studies among injured populations. It is important to note that spm1d analysis typically requires a larger sample size than discrete analysis. Consequently, many studies using spm1d may be underpowered and susceptible to type II errors (false negatives).^138^ In 2017, Pataky et al. introduced a tool to estimate the sample size for spm1d studies, but none of the studies in this review used it to calculate their sample size.^139^ This lack of methodological consistency may impact the comparability of study outcomes. To address these challenges, it is crucial to disseminate knowledge about the applications of spm1d and provide coding training to researchers within the sports science community. By increasing familiarity and understanding of spm1d’s capabilities, its adoption, and utilization are likely to broaden, leading to improved study design and more robust research findings in sports biomechanics.

Our findings revealed a diverse range of movements investigated using spm1d, both in healthy and injured populations. However, the utilization of spm1d varied across different movements and sports. Running, cutting, jumping/landing, and squatting were the most commonly examined activities among healthy participants, while running, soccer, and handball were the most frequently studied sports. This discrepancy highlights the need to explore other areas that have received less attention.

Most studies were conducted in controlled laboratory environments, leading to low ecological validity and may not fully replicate real-world conditions. With the emergence of wearable technologies, such as smart clothing, inertial sensing, and fitness trackers, researchers have the opportunity to extend the scope of applications to various sports and activities, incorporating more realistic conditions to represent real-world environments.

Additionally, our review underscored the imbalance between studies on healthy populations and those involving injured individuals. Future research on injured people could provide valuable insights into injury mechanisms, prevention, and rehabilitation strategies. While studies on ACL injuries, CAI, and hip-related pain were relatively common, other types of injuries, such as patellofemoral pain syndrome and hamstring injuries, common among athletes, have received less attention within the spm1d framework. Broadening the scope of research to encompass a broader range of injuries would enhance the utility of spm1d in sports medicine.

The included studies predominantly focused on adult populations, leaving gaps in our understanding of active pediatrics and older adults. Moreover, most of the studies did not focus specifically on gender. Considering the unique biomechanical profiles of different demographic and age groups, future studies should strive for inclusivity and account for these differences in their analyses. By extending the use of spm1d to these populations, our understanding of lower limb biomechanics across genders and the lifespan can be enriched.

While many studies in our review concentrated on footwear design and development, broader use of spm1d can inform sports equipment design and protective gear development. Evaluating the impact of different shoes, clothing, and protective gear on sports biomechanics can assist manufacturers in optimizing their designs for enhanced performance and injury prevention.

Several limitations should be considered in our study. Firstly, to make it more feasible, we have only included studies that evaluated lower limb movements in common sports injuries and sports activities. Secondly, we focused exclusively on English and peer-reviewed studies. Lastly, our review was limited to publications in peer-reviewed journals, potentially overlooking relevant works in grey literature.

## CONCLUSIONS

Our scoping review indicates a growing use of spm1d in sports biomechanics, particularly in assessing lower limb movements such as running, cutting, jumping, and squatting, as well as in conditions such as ACL injuries, CAI, and hip-related pain. A knowledge gap remains in underrepresented sports movements and diverse demographic groups.

## Data Availability

All data produced in the present study are available upon reasonable request to the authors

## Appendix 1. Search Strategy for the different Search Engines

### 1.1 Embase

’statistical parametric mapping’:ti,ab,kw AND (knee:ti,ab,kw OR hip:ti,ab,kw OR ankle:ti,ab,kw)

### 1.2 PubMed

statistical parametric mapping[Title/Abstract] AND (knee[Title/Abstract] OR hip[Title/Abstract] OR ankle[Title/Abstract])

### 1.3 ProQuest

noft(statistical parametric mapping AND (knee OR hip OR ankle)) Additional limits - Document type: Article; Language: English

### 1.4 Web of Science

((TI=(statistical parametric mapping AND (knee OR hip OR ankle))) OR AB=(statistical parametric mapping AND (knee OR hip OR ankle))) OR AK=(statistical parametric mapping AND (knee OR hip OR ankle))

## Appendix 2. SPM1d in populations with Other Sport-Related Injuries

| Study | Participants Details |  |  |  | SPM Assessment |  |  |  |
| --- | --- | --- | --- | --- | --- | --- | --- | --- |
|  | Number of Injured Participants | Number of Healthy Participants | Sport participation | Activity | Joints | Measurement Tools | Kinetic/ Kinematic | SPM/ Discreet |
| Houston (2021)<sup>136</sup> | 25 | 24 | Army | Single-leg squat, walking | Hip, Knee | Vicon + AMTI | Kinematics + Kinetics | SPM |
| Alizadeh (2022)<sup>104</sup> | 31 | 68 | Soccer | Isokinetic device | Knee | IsoMed 2000 | Kinetics | SPM+ Discreet |
| Pieters (2022)<sup>137</sup> | 56 | 60 | Soccer | Isokinetic device | Hip, Knee | Biodex | Kinetics | SPM |

## Notes

### Competing Interest Statement

The authors have declared no competing interest.

### Clinical Protocols

https://osf.io/aj32n

### Funding Statement

This study did not receive any funding

### Summary of Updates

- Fixed author last updated. - In-text references style update. - Two errors in paper numbers within the text are fixed.

